# The relationship between physical function and psychological symptoms in Parkinson’s: A Survey of People with Parkinson’s and Carers

**DOI:** 10.1101/2024.09.06.24313094

**Authors:** Philip Hodgson, Alastair Jordan, Charikleia Sinani, Divine Charura

**Author notes:** Corresponding Author Philip Hodgson, Physiotherapy, West Park Hospital, Edward Pease Way, Darlington DL2 2TS, United Kingdom.

## Abstract

**Background:** People with Parkinson’s (PwP) can experience both physical and psychological symptoms, and understanding the perspectives of people affected is crucial for improved management, and clinical outcomes.

**Objectives:** This online survey sought to investigate whether individuals perceive a connection between physical and psychological symptoms, while also considering the influence of personal roles and past symptom experiences.

**Methods:** A UK-wide survey of 251 PwP and 61 family/carers was conducted. The survey focused on reported diagnosed and non-diagnosed psychological symptoms experienced, their onset, and the perceived impact of physical and psychological symptoms on one another. Responses were summarised using descriptive statistics.

**Results:** A substantial proportion of respondents reported at least one diagnosed psychological condition (38.5%) or undiagnosed psychological symptoms (44.6%) such as anxiety and depression. Half of respondents reported perceiving a bi-directional interaction between physical and psychological symptoms, with this perception most reported in people with prior experience of psychological symptoms. Our sample shows that while PwP and carers have similar views on the impact of psychological symptoms, carers perceive the impact of physical symptoms to be greater than PwP.

**Conclusions:** PwP and carers appear to perceive an interaction between physical and psychological symptoms in Parkinson’s, noting that psychological symptoms frequently precede Parkinson’s diagnosis but are often under-recognised. Improved awareness of the potential link between physical and psychological symptoms in PwP may help to improve assessment, and onward referral processes to enhance care. Further research may assist in identifying potential sub-groups and allow the prediction of changes in physical and psychological presentation.

## 1. Introduction

In addition to common physical symptoms, Parkinson’s Disease (PD) can affect an individual’s mental wellbeing (1). People with Parkinson’s (PwP) experience higher rates of mental health issues, including depression, anxiety, schizophrenia, and psychotic symptoms, compared to the general population (1, 2). For example, whilst 17% of the general population will face anxiety and depression (3), this figure rises to 40% among PwP (4, 5). It is believed that this increased likelihood of mental health symptoms is linked to the condition itself or the side effects of medications (6). Despite these concerning statistics, current NICE guidelines (7) do not offer specific recommendations for addressing mental health issues in PwP. Instead, they simply refer to generic guidelines for depression in adults with chronic health conditions and suggest access to allied health professionals (AHP’s) such as physiotherapists, and PD nurse specialists. This approach contrasts with guidance for other neurological conditions, including Multiple Sclerosis, which incorporates specific recommendations for regular cognitive, emotional, and mental health screenings (8, 9).

While evidence in older populations suggests a link between physical and psychological presentations (10), there is limited research confirming such a relationship in the PD population. Available studies indicate that PwP perceive anxiety as a factor in amplifying their physical symptoms (11), including increased instances of freezing of gait (12). Several studies have suggested a correlation between increased anxiety and greater severity of motor symptoms, as measured by the Unified Parkinson’s Disease Rating Scale (UPDRS) (13–16). To our knowledge, this relationship has yet to be confirmed using more specific measures of physical function, such as balance and mobility assessments, or when considering other psychological symptoms associated with PD (17, 18). Confirming this relationship using alternative measures widely used by healthcare professionals is a vital step to progressing clinical practice, particularly given the aforementioned NICE guidelines. Based on the high proportion of individuals with PD affected by psychological symptoms, further research in this area could significantly enhance our understanding of the potential interaction between physical function and psychological symptoms, potentially leading to improved care strategies.

Prior to completing this work, our group conducted a systematic review (19) which highlighted that despite many studies routinely collecting data for both physical and psychological outcomes, only one study included in this review has explicitly examined the relationship between these outcomes (20). Furthermore, our exploratory meta-regression analysis of extracted baseline group-level mean data from previous studies suggested a trend for the physical ability of PwP to reduce as symptoms of depression increase. The single study explicitly investigating the relationship between physical and psychological outcomes was completed by Still et al. (20) and completed bivariate correlation analysis for HADS score against performance on the DGI outcome measure. No significant correlation was found between DGI performance and either overall HADS score (r=0.269), HADS-A (r=0.132), or HADS-D (r=0.239). Significant correlations were found between self-reported motor disability (MDS-UPDRS Part 2) and HADS score (r=0.624), HADS-A (r=0.536), and HADS-D (r=0.481). This supports the suggestion of a potential interaction between physical function and psychological symptoms in PD, however, given the lack of an identified significant correlation with clinician-rated measures of physical function, suggests a potential mismatch in this relationship between participant and clinician assessments. This work has provided the foundation for future investigations, including the investigation of the potential mismatch between clinician-rated and participants self-reported measures of physical function.

To date, no survey research has explored the perspectives of PwP and carers of PwP regarding the potential interaction between physical and psychological symptoms in PD. This represents a significant gap in our understanding of the lived experiences of those affected by PD. Current evidence lacks direct input from PwP and carers on their perceptions of symptom interactions, offers limited understanding of how these interactions are experienced in daily life, and provides insufficient data on how perspectives might differ between PwP and carers of PwP. Given these under-researched areas, our study aimed to provide a platform for PwP and carers of PwP to share their experiences and insights. Through an online survey, we collected views on potential interactions between physical and psychological symptoms, as well as information on how these interactions are experienced. Understanding these factors is crucial for informing future research directions and improving clinical service delivery in PD care.

## 2. Materials and Methods

### 2.1 Ethics

Ethical approval was given by the School of Science, Technology and Health Research Ethics Committee at York St John University (STHEC0067). Following provision of the participant information sheet, informed consent was obtained. This was completed electronically at the start of the survey with participants confirming “I meet the eligibility criteria and consent to completing the survey”. We confirm that we have read the Journal’s position on issues involved in ethical publication and affirm that this work is consistent with those guidelines.

### 2.2 Developing the Survey

To ensure that the survey considered priority areas, feedback on questions was sought. A draft survey was produced, and feedback provided by three volunteers recruited through Parkinson’s UK (one carer and two PwP). Volunteers reported that the research purpose was clear and that they supported the completion of research in this area. Language used within the survey and associated documentation was edited based on feedback received. Estimated timescales for completion were adjusted from 20-minutes to 30-minutes to reflect more accurately the time taken for PwP to complete the survey. Following feedback, the topic areas addressed included: Demographics, Physical activity, Mental health, Symptom interactions, and Treatments. A copy of the survey questions and flow is available in the supplementary material.

### 2.3 Recruitment and Procedures

The survey was open 24/11/2022-21/03/2024 via Qualtrics, an online survey and questionnaire tool. The survey was promoted via the Parkinson’s UK Research Support Network and associated Parkinson’s research interest groups. The Research Support Network has approximately 7,000 members (May 2022), the vast majority of whom are PwP and partners, family members and carers of those with the condition living in the UK. An email was sent to members of the research support network via a monthly e-newsletter, as well as being promoted via the Parkinson’s UK ‘Take Part Hub’.

The survey gathered responses from PwP, and family members or carers who were identified by response to the first survey question. No specific requests were made to identify people with specific physical and/or psychological symptoms. Carers were requested to answer each question based on their perception of the person they provide care for.

Whilst some demographic information was collected, the survey did not collect any personally identifiable information. Participants were able to skip any questions they did not wish to answer. A copy of the survey questions and flow is available in the supplementary material.

### 2.4 Data Analysis

Descriptive statistics were used to characterise the sample and data analysed using Microsoft Excel.

## 3. Results

A total of 312 responses were received, of which 251 were from PwP and 61 from family/carers. Responses to questions are detailed in the text and table/figures below. Please note that in some instances participants were able to select more than one response per question.

### 3.1 Participant Demographics

Table 1 shows details of participant demographics. Of the 312 respondents, 251 were from PwP, with the remainder from family/carers. PwP completing the survey tended to be aged 60 or over (74.1%), whilst family/carers were generally younger. Overall, 57.4% of respondents were female. For family/carer respondents, 90.2% were female, whilst this was 49.4% in responses from PwP. Responses were overwhelmingly from those identifying as white British (93.6%), with this being evident in both groups (PwP: 93.2%, family/carers: 95.1%). In PwP, time since PD diagnosis ranged from 0.01 years to 22.52 years, with a mean of 5.32 years. Time since PD diagnosis tended to be higher in responses from family/carers, with a mean of 10.56 years.

**Table 1:**
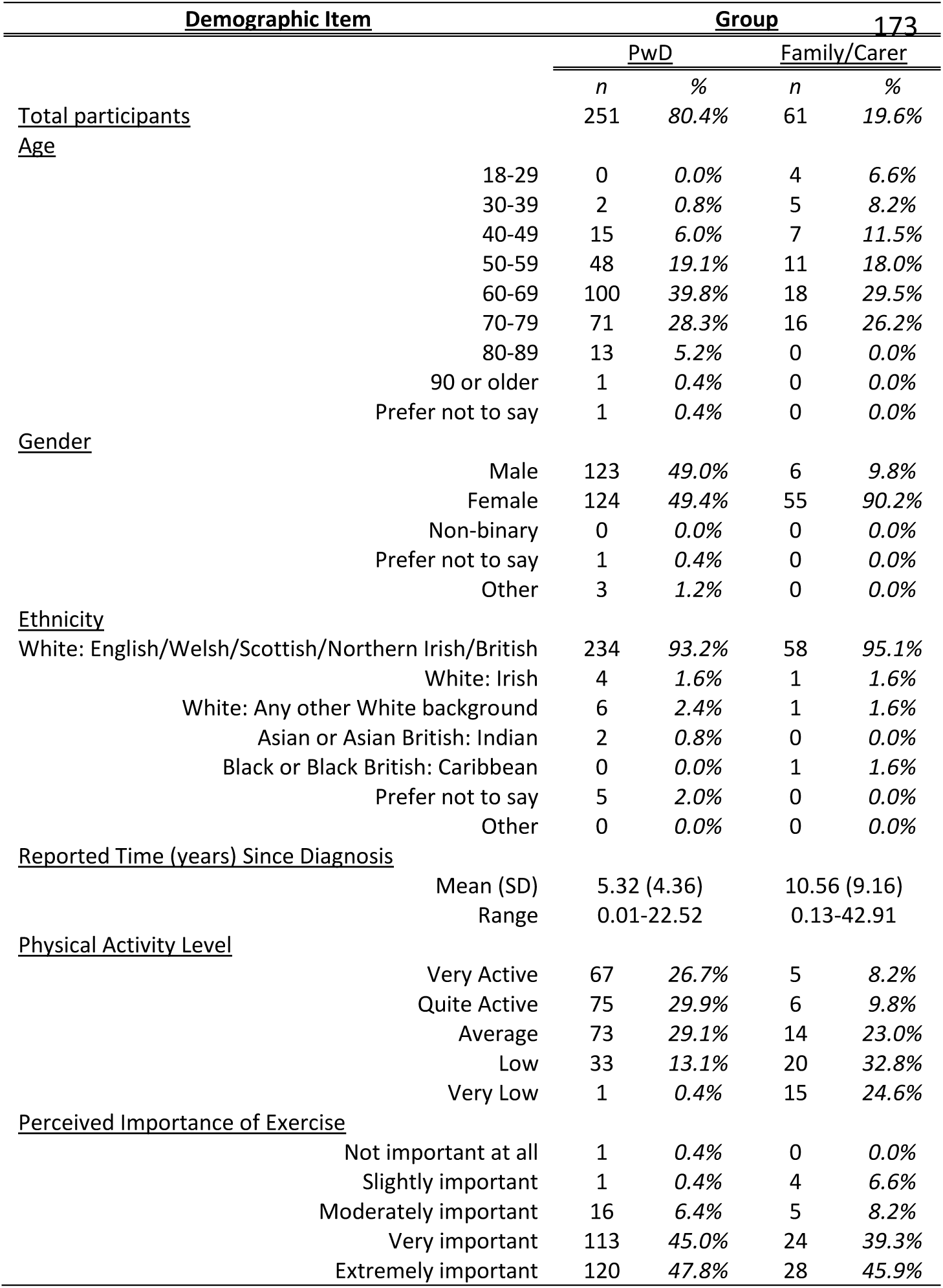
Participant demographics and physical activity level/importance

| Demographic Item |  | Group |  | 173 |  |
| --- | --- | --- | --- | --- | --- |
|  |  | PwD |  | Family/Carer |  |
|  |  | n | % | n | % |
| Total participants |  | 251 | 80.4% | 61 | 19.6% |
| <u>Age</u> |  |  |  |  |  |
|  | 18-29 | 0 | 0.0% | 4 | 6.6% |
|  | 30-39 | 2 | 0.8% | 5 | 8.2% |
|  | 40-49 | 15 | 6.0% | 7 | 11.5% |
|  | 50-59 | 48 | 19.1% | 11 | 18.0% |
|  | 60-69 | 100 | 39.8% | 18 | 29.5% |
|  | 70-79 | 71 | 28.3% | 16 | 26.2% |
|  | 80-89 | 13 | 5.2% | 0 | 0.0% |
|  | 90 or older | 1 | 0.4% | 0 | 0.0% |
|  | Prefer not to say | 1 | 0.4% | 0 | 0.0% |
| <u>Gender</u> |  |  |  |  |  |
|  | Male | 123 | 49.0% | 6 | 9.8% |
|  | Female | 124 | 49.4% | 55 | 90.2% |
|  | Non-binary | 0 | 0.0% | 0 | 0.0% |
|  | Prefer not to say | 1 | 0.4% | 0 | 0.0% |
|  | Other | 3 | 1.2% | 0 | 0.0% |
| <u>Ethnicity</u> |  |  |  |  |  |
|  | White: English/Welsh/Scottish/Northern Irish/British | 234 | 93.2% | 58 | 95.1% |
|  | White: Irish | 4 | 1.6% | 1 | 1.6% |
|  | White: Any other White background | 6 | 2.4% | 1 | 1.6% |
|  | Asian or Asian British: Indian | 2 | 0.8% | 0 | 0.0% |
|  | Black or Black British: Caribbean | 0 | 0.0% | 1 | 1.6% |
|  | Prefer not to say | 5 | 2.0% | 0 | 0.0% |
|  | Other | 0 | 0.0% | 0 | 0.0% |
| <u>Reported Time (years) Since Diagnosis</u> |  |  |  |  |  |
|  | Mean (SD) | 5.32 (4.36) |  | 10.56 (9.16) |  |
|  | Range | 0.01-22.52 |  | 0.13-42.91 |  |
| <u>Physical Activity Level</u> |  |  |  |  |  |
|  | Very Active | 67 | 26.7% | 5 | 8.2% |
|  | Quite Active | 75 | 29.9% | 6 | 9.8% |
|  | Average | 73 | 29.1% | 14 | 23.0% |
|  | Low | 33 | 13.1% | 20 | 32.8% |
|  | Very Low | 1 | 0.4% | 15 | 24.6% |
| <u>Perceived Importance of Exercise</u> |  |  |  |  |  |
|  | Not important at all | 1 | 0.4% | 0 | 0.0% |
|  | Slightly important | 1 | 0.4% | 4 | 6.6% |
|  | Moderately important | 16 | 6.4% | 5 | 8.2% |
|  | Very important | 113 | 45.0% | 24 | 39.3% |
|  | Extremely important | 120 | 47.8% | 28 | 45.9% |

### 3.2 Physical Activity

Table 1 also shows details of the reported physical activity levels, alongside details of the perceived importance of physical activity. The majority of responses from PwP indicate physical activity levels of average or higher (85.7%). Although family/carer responses do not necessarily relate to the same individuals, responses show a difference between the perception of the two groups, with only 41.0% of family members/carers reporting that the individual they care has activity levels of average or higher. Despite this mismatch in reported activity levels, both groups report physical activity to be either very important or extremally important (PwP: 92.8%, family/carers: 85.2%).

### 3.3 Psychological Symptoms and Diagnoses

#### 3.3.1 Presence of reported psychological diagnoses/symptoms, and frequency by condition

Figure 1 details reported formal psychological diagnoses received and symptoms reportedly experienced without receiving a formal diagnosis. In the PwP group, 79 (31.5%) reported having received a formal diagnosis of psychological condition(s). Of these, the most commonly reported were anxiety (19.1%) and depression (20.3%). A total of 41 family/carers (67.2%) reported that the individual they provide care for had received a formal diagnosis of psychological condition(s). Of the diagnoses reported by family members/carers, the most common were also anxiety (36.1%) and depression (37.7%).

**Figure 1:**
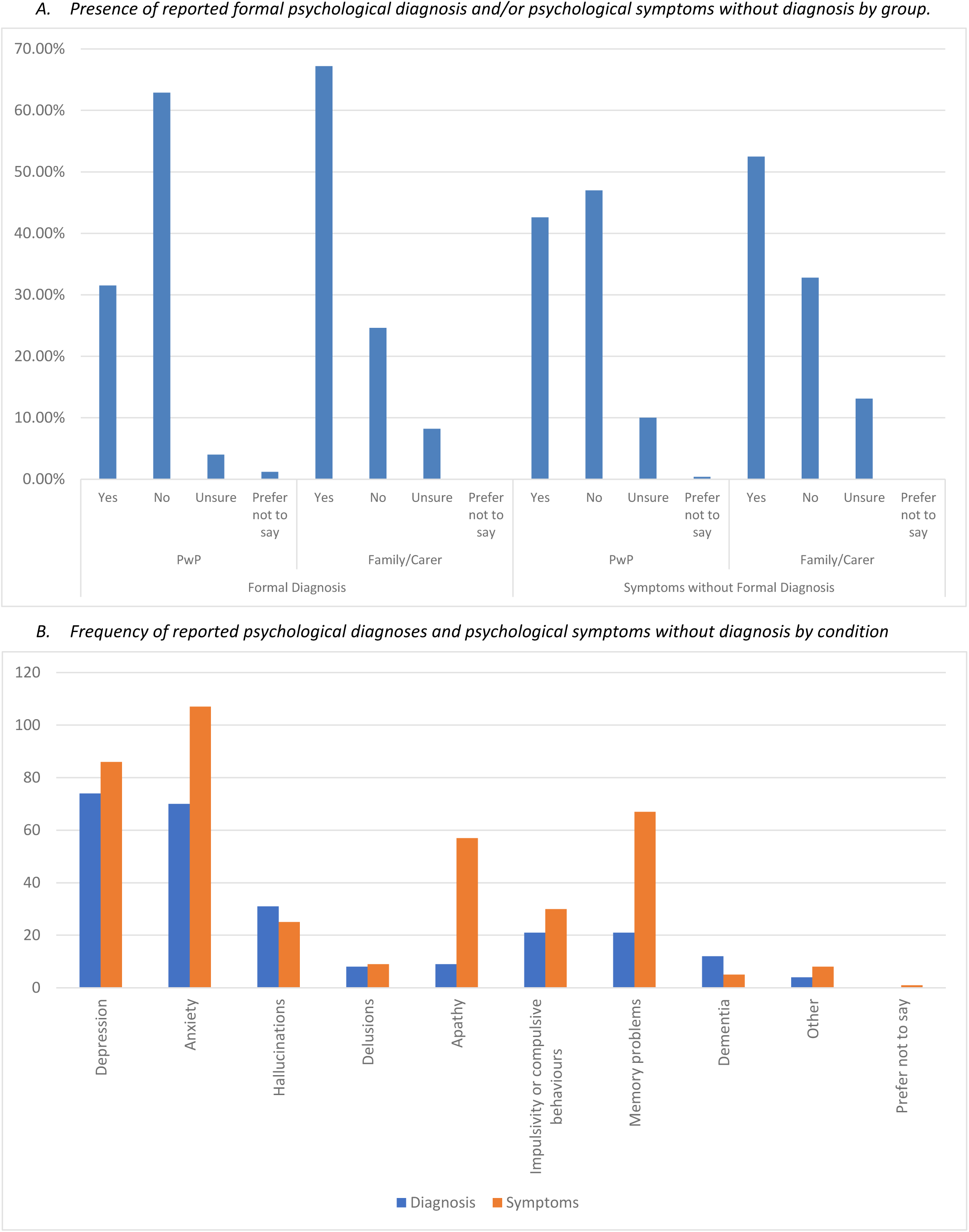
Details of psychological diagnoses and symptoms reported, and the frequency of reported psychological diagnoses and symptoms by condition

A greater proportion of PwP (107, 42.6%) reported having experienced symptoms of psychological condition(s) without receiving a formal diagnosis. The most commonly reported symptoms in this instance were anxiety (35.9%), depression (27.1%), apathy (17.1%) and memory problems (19.1%). A total of 32 family members and carers (52.5%) reported that the individual they provided care for had experienced symptoms of psychological conditions without receiving a formal diagnosis. The most commonly reported symptoms in this instance were also anxiety (27.9%), depression (29.5%), apathy (23.0%) and memory problems (31.1%). Psychological symptoms and diagnoses reported as ‘other’ included: ‘Post-Traumatic Stress Disorder’, ‘Bipolar Disorder’, and ‘Stress’, however accounted for 1.86% of responses to this question and have therefore not been considered further within data analysis.

#### 3.3.2 Timepoint of reported diagnoses/symptoms

Figure 2 expands on the reported diagnoses and symptoms to provide details around the timepoint at which the various diagnoses and symptoms were first given or experienced. This data takes into account the number of individuals reaching each timepoint based on the reported date of Parkinson’s diagnosis. From all responses received, the greatest likelihood of receiving a formal diagnosis of the following conditions was as follows: Depression – pre-PD diagnosis (63.5%), Anxiety – pre-PD diagnosis (55.7%), Hallucinations – Greater than 10y post-PD diagnosis (35.3%), and Impulsivity or Compulsive behaviours – 2-5y post-PD diagnosis (33.3%).

**Figure 2:**
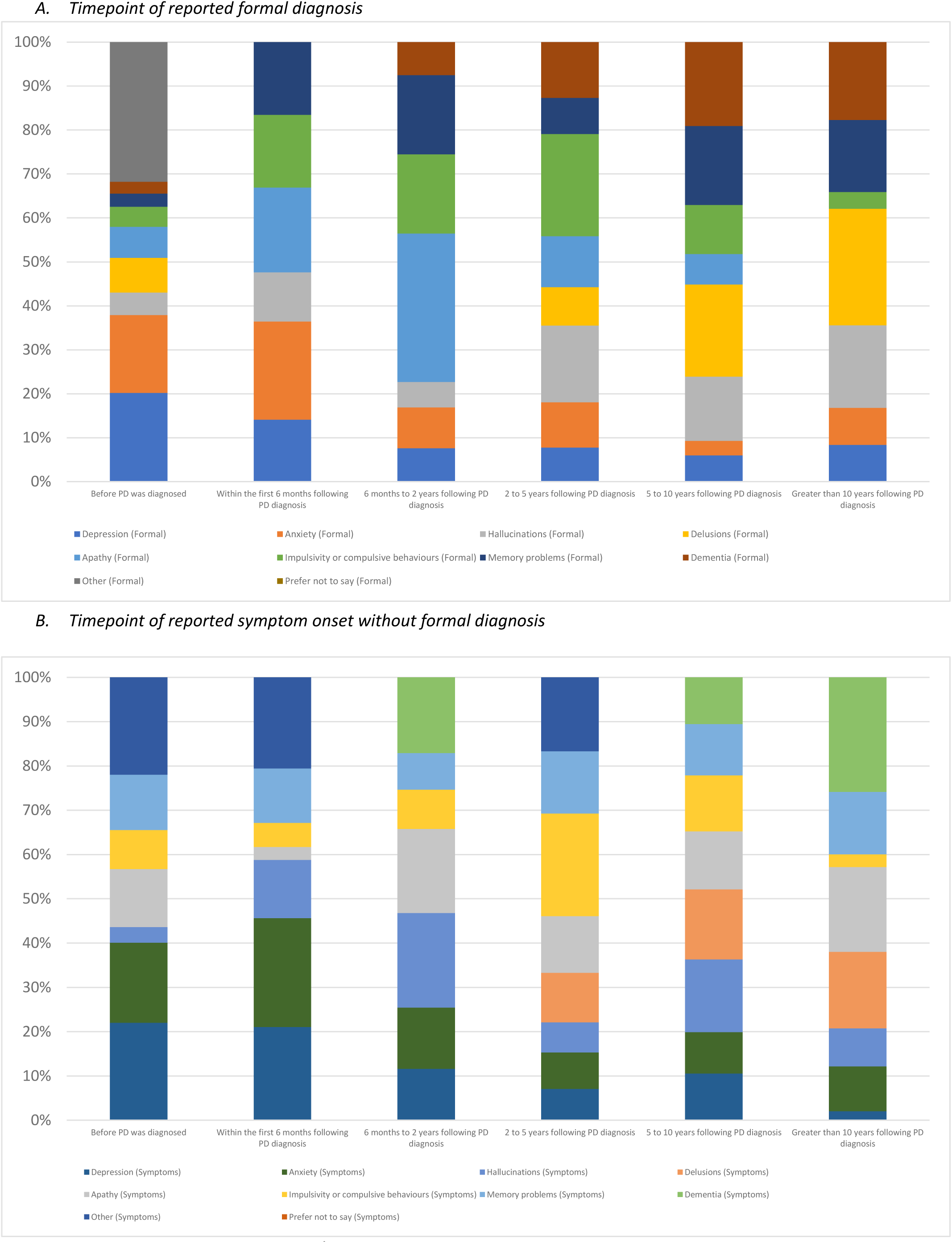
Timepoint of reported diagnoses/symptoms

From all responses received across both groups, the greatest likelihood of experiencing symptoms yet not receiving a formal diagnosis was as follows: Depression – pre-PD diagnosis (50.0%), Anxiety – pre-PD diagnosis (41.1%), Apathy – Greater than 10y post-PD diagnosis (55.6%), and Memory Problems – Greater than 10y post-PD diagnosis (40.9%). From our data, there appears to be greater variation in the reported timeframe of symptoms experienced without diagnosis in comparison to the timepoints reported for when formal diagnoses were received.

### 3.4 Perceived Symptom Interactions

Figure 3 shows details of reported symptom interactions as perceived by respondents. This data provides additional subgroup analysis for individuals reporting the presence of psychological symptoms and/or diagnosis in comparison to individuals not reporting these.

**Figure 3:**
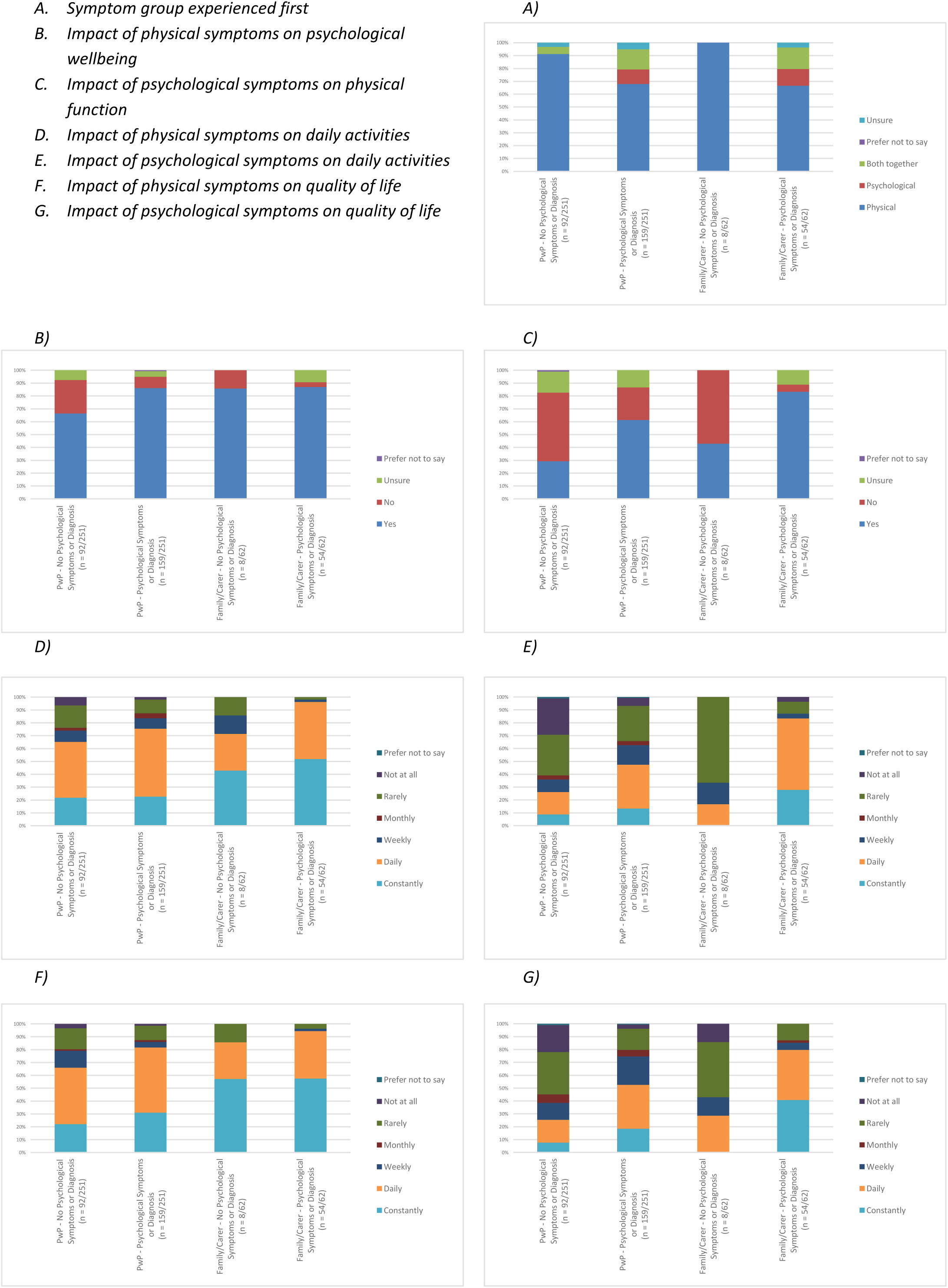
Perceived Symptom Interactions

#### 3.4.1 Symptoms Experienced First

Graph A shows that despite the high prevalence of psychological diagnoses and symptoms reported pre-PD diagnosis, the majority of PwP and family/carers report experiencing physical symptoms of PD prior to any psychological symptoms (76.5% and 70.5% respectively). This is higher in PwP and family/carers not reporting psychological symptoms (91.3% and 87.5%), dropping to 67.9% and 66.7% when considering only those who reported the presence of psychological symptoms and/or diagnosis.

#### 3.4.2 Broad Psychical and Psychological Symptom Interaction

Overall, both PwP and family/carer groups report perceiving physical symptoms to impact on psychological wellbeing (PwP: 78.9%, family/carers: 86.9%). This was again higher when considering PwP and carers who reported the presence of psychological symptoms and/or diagnosis (86.2% and 87.0% respectively) in comparison to those not reporting psychological symptoms and/or diagnosis (66.3% and 75.0%). When considering the inverse relationship, 49.4% of PwP and 78.7% of family/carers reported that they perceived psychological symptoms to impact upon physical function. Once again, this was higher in those reporting psychological symptoms/diagnosis (PwP: 61.0%, family/carers: 83.3%) when compared to PwP and family/carers not reporting psychological symptoms and/or diagnosis (29.3% and 37.5%). These results are shown in graphs B and C. Overall, 156 individuals (50.0%) reported perceiving that both physical and psychological symptoms impacted on one another. Of these, 127 (81.4%) were individuals with prior experience of psychological symptoms and/or diagnosis.

#### 3.4.3 Psychical and Psychological Symptom Interaction (Daily Activities and Quality of Life)

Follow-up questions were asked to ascertain the extent of any impact, with questions addressing the impact of both physical and psychological symptoms on daily activities (graphs D and E) and overall quality of life (graphs F and G). Responses indicate that both groups of PwP and family/carers perceive a ‘constant’ or ‘daily’ impact of physical symptoms on both daily activities (PwP: 71.7%, family/carers: 91.9%) and quality of life (PwP: 75.3%%, family/carers: 91.9%). Based on responses received, a lower impact is perceived on daily activities (PwP: 39.44%, family/carers: 74.2%) and quality of life (PwP: 42.2%%, family/carers: 72.6%) as a result of psychological symptoms. These views regarding the impact of psychological symptoms again appear impacted by the presence of reported psychological symptoms/diagnosis, with individuals reporting psychological symptoms/diagnosis perceiving a greater impact of psychological symptoms on daily activities (PwP: 47.2%, family/carers: 83.4%) and quality of life (PwP: 52.2%, family/carers: 79.6%) when compared to PwP and family/carers not reporting psychological symptoms/diagnosis (Daily activities: 26.1% and 12.5% and quality of life 25.0% and 25.0%).

## 4. Discussion

This online survey sought to investigate whether individuals perceive a connection between physical and psychological symptoms, while also considering the influence of personal roles and past symptom experiences. To our knowledge, this is the first study to explore the perspectives of PwP and family members/carers, which provides a direct account of how these issues are viewed by the recipients of clinical care. This work will help to provide a basis for service development alongside aiding the design of future clinical research projects.

Through the analysis of responses, we been able to consider the influence of individuals role and previous symptom experience on their perception of any psycho-physical symptom interactions. This research has helped to improve our understanding of how PwP and family/carers perceive the relationship between physical and psychological symptoms experienced, including at which timepoint various psychological symptoms are most likely to occur and which are most likely to be formally diagnosed.

### 4.1 Physical Activity

Previous research has shown that PwP generally perceive exercise as highly important in maintaining their level of physical function (21). This perspective is maintained within our study giving reassurance on the representativeness of our sample. When considering group differences, responses highlight a potential mismatch between PwP and carers in the reported levels of physical activity ultimately achieved. This may indicate a possible difference in opinion between PwP and carers in terms of what constitutes a low/high level of physical activity. Alternatively, this could indicate that carers completing our survey represent individuals experiencing a more significant physical and/or psychological impact of PD. This seems to be corroborated by carers reporting a longer time since diagnosis (10.56 years) compared to PwP (5.32 years), which may suggest additional physical decline (22). It is important however to note that the PwP and carer samples are not matched and do not necessarily represent different perspectives of the same individuals.

### 4.2 Psychological Symptoms and Diagnoses

#### 4.2.1 Presence of reported psychological diagnoses/symptoms, and frequency by condition

Participants were asked to recall the presence of formal psychological diagnoses and symptoms without formal diagnosis overall, before being asked to select the timepoint at which these diagnoses or symptoms were given/began. These timepoints included pre-Parkinson’s diagnosis, diagnosis to 6-months post-diagnosis, 6-months to 2-years post-diagnosis, 2-5 years post-diagnosis, and over 10-years post-diagnosis. These timepoints were chosen in order to capture information relating to significant events such as immediately following Parkinson’s diagnosis, but also capture changes potentially relating to disease progression and the impact of elements such as medication. Data analysis for the presence of symptoms/diagnosis was completed by group, whereas timepoint data was combined responses from PwP and family/carers to achieve a more balanced spread across the length of time since formal Parkinson’s diagnosis.

Since not all symptoms experienced result in or warrant a formal diagnosis, it is to be expected that some individuals will report symptoms without these having been acknowledged formally by clinicians. The reported rates of anxiety and depression in our survey appear to agree with the previously reported prevalence rates (5). Carers reported higher rates of psychological symptoms and diagnoses beyond anxiety and depression. This trend may be attributed to the longer duration since diagnosis in the carer group, allowing more time for various symptoms to manifest and be formally identified.

#### 4.2.2 Timepoint of reported diagnoses/symptoms

In our survey, anxiety and depression were most likely to be reported prior to formal diagnosis of Parkinson’s. This is of interest given that an increase in symptoms of depression is thought to precede PD diagnosis by a few years (23), with a similar proposal for anxiety (24). It is impossible to say with certainty whether the presence of anxiety and depression prior to formal diagnosis of Parkinson’s represents the presence of Parkinson’s itself at this stage, however the occurrence of anxiety and depression is higher in PwP than in the general population (25). It is also important to note that it is difficult to fully interpret such findings due to the complexity of any potential interactions, particularly as any relationship between physical and psychological symptoms is likely to be bi-directional in nature.

The temporal relationship between the onset of anxiety and depression and the formal diagnosis of PD warrants further investigation. It is possible that the neurodegenerative processes underlying PD contribute to the development of anxiety and depression, or conversely, that these psychological conditions exacerbate the physical symptoms of PD. Additionally, the stress and uncertainty associated with the early, undiagnosed stages of PD could also play a role in the emergence of these symptoms. Future research should aim to disentangle these complex interactions to better understand the relationship between physical and psychological symptoms in PD, and to develop more effective strategies for early diagnosis and comprehensive treatment.

Symptoms of apathy and impulsivity were also commonly reported within our sample; however, both were more likely to be experienced as symptoms rather than involving formal recognition. Symptoms of apathy were commonly reported throughout the disease time-course, with symptoms of impulsivity also reported throughout however appeared to peak at 2-5 years post Parkinson’s diagnosis, consistent with previous findings (26). In our sample, symptoms of hallucinations were most commonly reported 2-5 years following Parkinson’s diagnosis, with formal diagnosis peaking at over 10 years following Parkinson’s diagnosis, likely reflecting the side-effects of levodopa treatment and limited treatment for the hallucinations themselves without inducing further side-effects (27).

These findings have implications for clinical services with respect to screening for various psychological symptoms and subsequent education provided following diagnosis. For example, given the high reported prevalence of anxiety and depression symptoms prior to formal Parkinson’s diagnosis, it may be beneficial for clinical services to screen for these symptoms from the outset to enable optimal signposting to relevant secondary services. Whilst not a conclusive resource for Parkinson’s symptoms at each timepoint, this work provides some insight into self-reported symptoms which are not necessarily identified by clinical services. In addition, this work provides a platform for further investigation regarding the presence of various symptoms alongside actions taken on identification within practice. As others have suggested, it may eventually be possible to predict the presence or likelihood of developing Parkinson’s based on the identification of non-motor symptoms prior to the development of observable motor symptoms (28–31).

### 4.3 Perceived Symptom Interactions

#### 4.3.1 Symptoms Experienced First

As discussed previously, there is evidence to suggest that non-motor symptoms may precede diagnosis of PD, and even the motor symptoms themselves (23). It is therefore somewhat surprising that despite many respondents reporting non-motor symptoms/diagnosis prior to diagnosis of PD, over 75% nevertheless report experiencing motor symptoms first. This may indicate a lengthy process of diagnosis in which both groups of symptoms are experienced before formal diagnosis, a recall bias towards motor symptoms, a lack of recognition that non-motor symptoms may be associated with PD even amongst individuals directly impacted by the condition, or pre-existing non-motor symptoms long before the presence of PD was considered.

#### 4.3.2 Broad Psychical and Psychological Symptom Interaction

Individuals who had reported psychological symptoms and/or a formal diagnosis reported an increased recognition of the potential for psychological symptoms to occur prior to physical symptoms. In comparison to individuals without experience of psychological symptoms, this group were more likely to perceive an interaction between physical and psychological symptom on one another. Strikingly, although 50.0% of participants overall reported perceiving that both physical and psychological symptoms impacted on one another, 81.4%) of these were individuals with prior experience of psychological symptoms and/or diagnosis. This may reflect an under-recognition of potential symptom interactions until directly experienced by an individual, and potentially highlight a gap in education from clinicians when preparing people for what to expect as their condition progresses (32, 33). This may be particularly relevant when preparing for the later stages of the condition, or when explaining to individuals that the psychological symptoms they have experienced prior to formal PD diagnosis may be related to the presence of PD itself.

This latency in the awareness of the broader impact of physical and psychological symptoms is also reflected in the responses to other questions, suggesting a direct impact of personal experiences (i.e. the presence or absence of psychological symptoms) and an individual’s role (PwP or family/carer) on a person’s perception. The primary factor involved with this appears to be whether the question is concerned with the impact of physical or psychological symptoms.

#### 4.3.3 Psychical and Psychological Symptom Interaction (Daily Activities and Quality of Life)

Based on responses to our survey, the presence or absence of psychological symptoms tends to have a greater impact on how individuals perceive psychological symptoms to impact on aspects such as physical function, daily activities, and quality of life. Individuals without experience of psychological symptoms were much less likely to report a perceived impact of psychological symptoms in these areas in comparison to individuals with first-hand experience of psychological symptoms. These responses might reinforce our suggestion that there is an under-recognition of symptom interactions until directly experienced and suggests a role for clinicians in preparing for these scenarios in advance.

In addition to the impact of personal experiences, there also appears to be an impact of an individual’s role in the response to these questions, with PwP and family/carers responding differently to questions concerning the influence of physical symptoms regardless of their experience’s psychological symptoms. In general, responses from family/carers reflect a greater perceived impact of physical symptoms on daily activities and quality of life in comparison to PwP. This was initially surprising however when given further consideration may be because carers are potentially not present at all times and may therefore more likely to overestimate the impact of these symptoms in comparison to PwP, who may themselves have simply become more accustomed to this impact and adapted their lifestyle accordingly. Alternatively, as family/carer responders do not necessarily reflect the same individuals with Parkinson’s, and tended to report a longer time since diagnosis, this difference may merely signal an influence of greater symptom progression.

### 4.4 Strengths and Limitations of this study

Whilst efforts were made to minimise limitations, this study lacked the ability to objectively assess disease severity and compare this with reported symptoms. Although our survey collected a large number of responses, indicating that many individuals felt strongly enough about the topic to complete it, the sample was non-random. As a result, respondents were potentially more likely to have experienced both physical and psychological symptoms or have an interest in this area, so may not be entirely representative of the Parkinson’s population overall. Given the immense variety of potential symptoms that individuals may experience, information was not collected regarding the specific physical symptoms, or the severity of symptoms that were experienced by PwP completing our survey. Due to the subjective nature of survey responses, it is therefore not possible to infer any interactions between specific physical and psychological symptoms, however this is a potential opportunity for further investigation now that an overall perspective has been established.

Where possible we have attempted to compare our data to previously published research as a reference point to indicate the representativeness of our sample. For example, our sample reported a high perceived importance of physical activity, and the reported rates of anxiety and depression aligned with the accepted prevalence in PD. Respondents were all from the United Kingdom, which has implications for the availability of clinical services. Despite these limitations, our survey was able to address the aim of this exploratory study, to provide an opportunity for PwP and carers of PwP to share their experiences of any potential interaction between physical and psychological symptoms experienced.

A major strength of this study was that it allowed the exploration of the perceived relationship between physical and psychological symptoms of Parkinson’s by collecting the views of PwP and carers. PwP and carers themselves were involved in the design of the survey through commenting on draft questions to ensure that the survey focused on areas that were most important to them. The work has helped to provide an overview of how any relationship between these symptom groups is perceived, which will be helpful when combined with ongoing work considering the perspectives of UK-based physiotherapists. The study highlights the need to enhance formal assessment of psychological symptoms and optimising onward referrals to appropriate services to provide comprehensive care for PwP.

## 5. Conclusions

From our survey results, PwP and carers appear to appreciate the link between physical function and non-motor symptoms in Parkinson’s, however both report an under-recognition of psychological symptoms experienced. Anxiety and depression are most commonly reported as being experienced and formally diagnosed, yet there is large variation in the timepoint of reported diagnosis and symptoms. Psychological symptoms were widely reported to precede formal diagnosis of Parkinson’s in many cases. Furthermore, the extent of these reported perceived symptom interactions, and the wider impact of each, appears to be influenced both by role (PwP or family/carer) and by whether an individual has first-hand experience of psychological symptoms.

The findings of this study highlight the complex interplay between physical and non-motor symptoms in Parkinson’s disease, emphasising the importance of a holistic approach to patient care. The perceived bi-directional relationship between these symptom groups, particularly among those with prior experience of psychological symptoms, underscores the need for comprehensive assessment and management strategies. The discrepancy in symptom impact perception between people with Parkinson’s and carers further emphasises the importance of considering multiple perspectives in care planning. By recognising the interconnectedness of symptom groups, healthcare providers can develop more effective, personalised treatment plans that address multiple aspects of the disease, potentially improving overall quality of life for individuals living with Parkinson’s.

Given the widely reported perceived interaction between physical and psychological symptoms, efforts should be made to improve formal symptom recognition and optimise signposting to appropriate services. Enhancing formal assessment of psychological symptoms and optimising onward referrals to appropriate services is essential to provide comprehensive care for PwP. Further research is needed to identify any potential sub-groups and objectively predict the decline in physical function and psychological symptoms.

## 8. Supplementary Material

Supplemental material for this article is available online [DOI: 10.25421/yorksj.25974733].

*This work was completed as part of an ongoing PhD being undertaken by the first author through a partnership between Tees, Esk and Wear Valleys NHS Foundation Trust and York St John University. The author(s) received no financial support for the research, authorship, and/or publication of this article.*

*Ethical approval was given by the School of Science, Technology and Health Research Ethics Committee at York St John University (STHEC0067). Informed consent was obtained electronically at the start of the survey, following provision of the participant information sheet.*

## Supporting information

Survey Questions and Flow

## Data Availability

Supplemental material for this article is available online [DOI: 10.25421/yorksj.25974733].

