## Supplementary material for "The relationship between physical function and psychological symptoms in Parkinson’s: A Survey of People with Parkinson’s and Carers": Survey Questions and Flow

Final Survey: 23/11/2022

**Appendix 2 – Survey Questions:**

### Understanding the relationship between Physical and Psychological Symptoms in Parkinson's Disease

### Survey Flow

**Standard: Welcome (1 Question)**
**Block: Informed Consent and group selection (3 Questions)**
**Standard: About You (4 Questions)**
**Standard: Physical Activity and Exercise (3 Questions)**
**Standard: About your Mental Health (23 Questions)**
**Standard: Impact of physical symptoms on Mental Health (3 Questions)**
**Standard: Impact of psychological symptoms on physical wellbeing: (3 Questions)**
**Standard: Treatments (5 Questions)**
**Standard: Covid-19 (5 Questions)**
**Standard: Mattering and Anti-Mattering (2 Questions)**
**Standard: Any further comments (1 Question)**

Page Break

Start of Block: Welcome

Welcome

**Understanding the relationship between Physical and Psychological Symptoms: Survey of People with Parkinson's Disease and their carers**

Individuals with Parkinson's Disease commonly experience psychological symptoms alongside the physical symptoms of their condition. At the moment we don't know much about the potential interaction between these physical and psychological symptoms in Parkinson's Disease. This study aims to explore the thoughts of people living with Parkinson's Disease and their carers about the relationship between the physical and psychological symptoms of Parkinson's Disease.

As a person living with Parkinson's Disease, or a person caring for people/person(s) with Parkinson's Disease, you have a unique understanding of the symptoms experienced and their impact on daily life. In order to improve our understanding of any relationship between the physical and psychological symptoms of Parkinson's Disease we invite you to complete this 30-minute survey exploring your experiences of common physical and psychological symptoms. Please read the following information carefully. If anything is unclear, please contact via the details at the bottom of this sheet.

If required, you can complete the survey in separate sessions, but will need to use the same device and browser to make sure your previous answers are stored. Once started, you will have 2-weeks to complete the survey.

To take part you must meet all of the following criteria:

1. Either be diagnosed with Parkinson's Disease or be a formal/informal carer for people/person(s) with Parkinson's Disease
2. Be aged over 18
3. Reside in the UK
4. Be fluent in English
5. Not have any cognitive impairments that impacts on your capacity to take part, for example your ability to read, understand, or answer questions

Taking part in this study involves completing a short survey about your experiences of common physical and psychological symptoms of Parkinson's Disease. There are a total of 37 questions, however you may skip any questions that you don't feel comfortable answering. Questions are mainly multiple choice however some questions invite additional comment should you wish to. We ask that you to take your time responding to the questions, and to please forward the link of the survey to other people living with Parkinson's Disease and/or carers of people living with Parkinson's Disease.

Final Survey: 23/11/2022

It is not felt that there is any direct risk to you from completing this survey. The survey might ask questions about topics that you may find sensitive. This information is important for us to truly understand your experience. If at any point you do not wish to continue with the survey, you can stop immediately. Should you require any physical or psychological support following completion of this survey please contact your local GP or relevant charity organisations such as Parkinson's UK via email at or telephone 0808 800 0303 (helpline) and MIND via email at or telephone 0300 123 3393 (helpline).

Your answers to the survey questions will be used for analysis and possible publication. The data will be stored in the York St John One Drive secure system. People will use this information to do the research or to make sure that the research is being done properly. We will keep all information about you safe and secure. Data will be disposed according to the UK Data Archive 'Managing and Sharing Data - best practice for researchers' guide (3rd edition). This document can be found here: https://ukdataservice.ac.uk/media/622417/managingsharing.pdf

This project is being completed as part of PhD research project undertaken by Philip Hodgson. Philip is an NHS Physiotherapist completing this research part-time alongside his clinical work. The project is supervised by Professor Divine Charura, Dr Alastair Jordan, and Dr Charikleia Sinani based at York St John University. Please contact Philip via email should you have any questions before completing the survey.

Approval to conduct this research has been provided by York St John University (INSERT REF), in accordance with its ethics review and approval procedures. Any person considering participation in this research project, or agreeing to participate, may raise any questions or issues with the researchers at any time. In addition, any person not satisfied with the response of researchers may raise ethics issues or concerns and may make any complaints about this research project by contacting Dr Charlotte Haines-Lyon, Chair of the Ethics Committee for the School of Education, Language and Psychology'. All research participants are entitled to retain a copy of any Participant Information Form and/or Participant Consent Form relating to this research project (please click here to download a copy).

Should you have any concerns relating to GDPR please use the following contact details: University Secretary, York St John University, Lord Mayor's Walk, York, YO31 7EX.

**Thank you in advance for your participation**

End of Block: Welcome

---

Final Survey: 23/11/2022

Start of Block: Informed Consent and group selection

**By clicking the 'agree' button you are providing consent to participate, in full knowledge of the information in the participant information sheet and agree with all the statements below:**

- I understand the contents of the Participant Information Sheet
- I have been given the opportunity to ask questions about the study and have had them answered satisfactorily.
- I understand that my participation is entirely voluntary, and I am under no obligation to complete this survey.
- I understand who will have access to my data, how it will be stored, in what form it will be shared, and what will happen to it at the end of the study.
- I understand that once I begin this survey my data will be anonymised and cannot then be withdrawn.
- I understand that data from incomplete surveys will be included in analysis where appropriate.
- I agree to take part in the study.

**All research participants are entitled to retain a copy of any Participant Information Form and/or Participant Consent Form relating to this research project (please click** here **to download a copy).**

○ I meet the eligibility criteria and consent to completing the survey

○ I do not consent or do not meet the eligibility criteria

*Skip To: End of Survey If Q1 = I do not consent or do not meet the eligibility criteria*

Page Break

Q1 Please select whether you are an individual diagnosed with Parkinson's Disease, or an individual providing care for individual(s) with Parkinson's Disease.

◯ I have been diagnosed with Parkinson's Disease

◯ I provide care for an individual or individuals with Parkinson's Disease

---

*Display This Question:*

*If Q1 = I provide care for an individual or individuals with Parkinson's Disease*

Q2 Please select which category most accurately describes the care you provide.

◯ Formal/Paid Carer

◯ Informal/Friend/Family Carer

◯ Prefer not to say

**End of Block: Informed Consent and group selection**

Start of Block: About You

Q3 What is your current age?

- ○ 18 - 29
- ○ 30 - 39
- ○ 40 - 49
- ○ 50 - 59
- ○ 60 - 69
- ○ 70 - 79
- ○ 80 - 89
- ○ 90 or older
- ○ Prefer not to say

Q4 What is your ethnicity?

○ White: English/Welsh/Scottish/Northern Irish/British

○ White: Irish

○ White: Any other White background

○ Gypsy or Irish Traveller

○ Asian or Asian British: Indian

○ Asian or Asian British: Pakistani

○ Asian or Asian British: Bangladeshi

○ Asian or Asian British: Any other Asian background

○ Chinese

○ Black or Black British: African

○ Black or Black British: Caribbean

○ Black or Black British: Any other Black background

○ Mixed: White and Black Caribbean

○ Mixed: White and Black African

○ Mixed: White and Asian

○ Mixed: Any other Mixed background

○ Arab

○ Prefer not to say

○ Other, please specify: ________________________________________________

Q5 What is your gender?

○ Male

○ Female

○ Non-binary

○ Prefer not to say

○ Other, please specify: ________________________________________________

---

*

Q6 Roughly when did you receive a formal diagnosis of Parkinson's Disease?

*If caring for an individual or individuals with Parkinson's Disease please enter an approximate earliest date at which their Parkinson's disease was formally diagnosed.*

Dates need to be in the format DD/MM/YYYY, for example 25/02/1992

________________________________________________________________

End of Block: About You

---

Start of Block: Physical Activity and Exercise

Q7 Which of the following best describes you:

*If completing as a carer for individual(s) with Parkinson's Disease please complete based on your perception of those you care for.*

- ○ Very active: Moderate to vigorous exercise for at least 150 minutes /week (eg 30 minutes five times a week). Moderate to vigorous exercise includes activities such as tennis, brisk walking, jogging, cycling, swimming.
- ○ Quite active: Moderate to vigorous exercise but for less than 150 minutes/week. Moderate to vigorous exercise includes activities such as tennis, brisk walking, jogging, cycling, swimming.
- ○ Average: Gentle exercise such as walking round the shops, bowls, fishing, golf (with a cart) and maintaining day-to-day independence.
- ○ Low: Active around the house but don't go out much.
- ○ Very low: Depend on others to help get about.

---

Q8 How important do you feel taking regular exercise is to keeping well with your Parkinson's disease?

*If completing as a carer for individual(s) with Parkinson's Disease please complete based on your perception regular exercise and keeping well with Parkinson's Disease.*

- ○ Not at all important
- ○ Slightly important
- ○ Moderately important
- ○ Very important
- ○ Extremely important

Q9 If you are not as active as you would like to be, what do you feel prevents you from being more active? Select all that apply.

*If completing as a carer for individual(s) with Parkinson's Disease please complete based on your perception of their barriers to physical activity for those you care for.*

- ☐ There is not enough time to exercise
- ☐ I am afraid of falling
- ☐ Fatigue - I feel exhausted all the time
- ☐ Apathy - I can't find the motivation to exercise
- ☐ Anxiety
- ☐ Depression
- ☐ Lack of confidence
- ☐ I don't have an exercise partner
- ☐ I am in poor general physical health
- ☐ It is too difficult to attend classes – either due to lack of transport or lack of someone to help me get there
- ☐ I prefer to exercise outdoors but the weather is too bad
- ☐ There are no suitable exercise classes
- ☐ My Parkinson's symptoms make it difficult to exercise (muscle stiffness, slow movement, shakiness)
- ☐ I don't believe exercise will help me
- ☐ I have to shield because of COVID-19

▢ Restrictions on socialising (due to COVID-19) limit my exercise options

▢ Other, please specify: ________________________________________________

▢ ⊗Nothing

End of Block: Physical Activity and Exercise

---

Start of Block: About your Mental Health

Q10 Have you ever been given a formal diagnosis of any psychological conditions by a healthcare professional?

*Examples may include (but are not limited to): Depression, Anxiety, Hallucinations, Delusions, Apathy, Impulsivity or compulsive behaviours, Memory problems, Dementia.*
*If completing as a carer for individual(s) with Parkinson's Disease please complete based on your knowledge of those you care for.*

◯ Yes

◯ No

◯ Unsure

◯ Prefer not to say

*Skip To: Q13 If Q10 != Yes*

Q11 If yes, what was this condition/conditions? Please select all that apply:

*If completing as a carer for individual(s) with Parkinson's Disease please complete based on your knowledge of those you care for.*

- ☐ Depression
- ☐ Anxiety
- ☐ Hallucinations
- ☐ Delusions
- ☐ Apathy
- ☐ Impulsivity or compulsive behaviours
- ☐ Memory problems
- ☐ Dementia
- ☐ Other, please specify: ________________________________________________
- ☐ ⊗Prefer not to say

*Display This Question:*

*If Q11 = Depression*

Q12a In a previous question you reported having been given a diagnosis of depression. Approximately, when were you given this diagnosis? *If completing as a carer for individual(s) with Parkinson's Disease please complete based on your knowledge of those you care for.*

- ○ Before Parkinson's Disease was diagnosed
- ○ Within the first 6 months following Parkinson's Disease diagnosis
- ○ 6 months to 2 years following Parkinson's Disease diagnosis
- ○ 2 to 5 years following Parkinson's Disease diagnosis
- ○ 5 to 10 years following Parkinson's Disease diagnosis
- ○ Greater than 10 years following Parkinson's Disease diagnosis
- ○ Unsure
- ○ Prefer not to say

*Display This Question:*

*If Q11 = Anxiety*

Q12b In a previous question you reported having been given a diagnosis of anxiety. Approximately, when were you given this diagnosis?

*If completing as a carer for individual(s) with Parkinson's Disease please complete based on your knowledge of those you care for.*

- ○ Before Parkinson's Disease was diagnosed
- ○ Within the first 6 months following Parkinson's Disease diagnosis
- ○ 6 months to 2 years following Parkinson's Disease diagnosis
- ○ 2 to 5 years following Parkinson's Disease diagnosis
- ○ 5 to 10 years following Parkinson's Disease diagnosis
- ○ Greater than 10 years following Parkinson's Disease diagnosis
- ○ Unsure
- ○ Prefer not to say

*Display This Question:*

*If Q11 = Hallucinations*

Q12c In a previous question you reported having been given a diagnosis of hallucinations. Approximately, when were you given this diagnosis?

*If completing as a carer for individual(s) with Parkinson's Disease please complete based on your knowledge of those you care for.*Click to write the question text

- ○ Before Parkinson's Disease was diagnosed
- ○ Within the first 6 months following Parkinson's Disease diagnosis
- ○ 6 months to 2 years following Parkinson's Disease diagnosis
- ○ 2 to 5 years following Parkinson's Disease diagnosis
- ○ 5 to 10 years following Parkinson's Disease diagnosis
- ○ Greater than 10 years following Parkinson's Disease diagnosis
- ○ Unsure
- ○ Prefer not to say

*Display This Question:*

*If Q11 = Delusions*

Q12d In a previous question you reported having been given a diagnosis of delusions. Approximately, when were you given this diagnosis?

*If completing as a carer for individual(s) with Parkinson's Disease please complete based on your knowledge of those you care for.*

- ◯ Before Parkinson’s Disease was diagnosed
- ◯ Within the first 6 months following Parkinson's Disease diagnosis
- ◯ 6 months to 2 years following Parkinson's Disease diagnosis
- ◯ 2 to 5 years following Parkinson's Disease diagnosis
- ◯ 5 to 10 years following Parkinson's Disease diagnosis
- ◯ Greater than 10 years following Parkinson's Disease diagnosis
- ◯ Unsure
- ◯ Prefer not to say

*Display This Question:*

*If Q11 = Apathy*

Q12e In a previous question you reported having been given a diagnosis of apathy. Approximately, when were you given this diagnosis?

*If completing as a carer for individual(s) with Parkinson's Disease please complete based on your knowledge of those you care for.*

- ○ Before Parkinson's Disease was diagnosed
- ○ Within the first 6 months following Parkinson's Disease diagnosis
- ○ 6 months to 2 years following Parkinson's Disease diagnosis
- ○ 2 to 5 years following Parkinson's Disease diagnosis
- ○ 5 to 10 years following Parkinson's Disease diagnosis
- ○ Greater than 10 years following Parkinson's Disease diagnosis
- ○ Unsure
- ○ Prefer not to say

*Display This Question:*

*If Q11 = Impulsivity or compulsive behaviours*

Q12f In a previous question you reported having been given a diagnosis of impulsivity or compulsive behaviours. Approximately, when were you given this diagnosis?

*If completing as a carer for individual(s) with Parkinson's Disease please complete based on your knowledge of those you care for.*

- ◯ Before Parkinson's Disease was diagnosed
- ◯ Within the first 6 months following Parkinson's Disease diagnosis
- ◯ 6 months to 2 years following Parkinson's Disease diagnosis
- ◯ 2 to 5 years following Parkinson's Disease diagnosis
- ◯ 5 to 10 years following Parkinson's Disease diagnosis
- ◯ Greater than 10 years following Parkinson's Disease diagnosis
- ◯ Unsure
- ◯ Prefer not to say

*Display This Question:*

*If Q11 = Memory problems*

Q12g In a previous question you reported having been given a diagnosis of memory problems. Approximately, when were you given this diagnosis?

*If completing as a carer for individual(s) with Parkinson's Disease please complete based on your knowledge of those you care for.*

- ◯ Before Parkinson's Disease was diagnosed
- ◯ Within the first 6 months following Parkinson's Disease diagnosis
- ◯ 6 months to 2 years following Parkinson's Disease diagnosis
- ◯ 2 to 5 years following Parkinson's Disease diagnosis
- ◯ 5 to 10 years following Parkinson's Disease diagnosis
- ◯ Greater than 10 years following Parkinson's Disease diagnosis
- ◯ Unsure
- ◯ Prefer not to say

*Display This Question:*

*If Q11 = Dementia*

Q12h In a previous question you reported having been given a diagnosis of dementia. Approximately, when were you given this diagnosis?

*If completing as a carer for individual(s) with Parkinson's Disease please complete based on your knowledge of those you care for.*

- ○ Before Parkinson's Disease was diagnosed
- ○ Within the first 6 months following Parkinson's Disease diagnosis
- ○ 6 months to 2 years following Parkinson's Disease diagnosis
- ○ 2 to 5 years following Parkinson's Disease diagnosis
- ○ 5 to 10 years following Parkinson's Disease diagnosis
- ○ Greater than 10 years following Parkinson's Disease diagnosis
- ○ Unsure
- ○ Prefer not to say

*Display This Question:*

*If Q11 = Other, please specify:*

Q12i In a previous question you reported having been given a diagnosis of 'Other' psychological condition(s). Approximately, when were you given this/these diagnosis? If multiple diagnoses please refer to the earliest date.

*If completing as a carer for individual(s) with Parkinson's Disease please complete based on your knowledge of those you care for.*

- ○ Before Parkinson's Disease was diagnosed
- ○ Within the first 6 months following Parkinson's Disease diagnosis
- ○ 6 months to 2 years following Parkinson's Disease diagnosis
- ○ 2 to 5 years following Parkinson's Disease diagnosis
- ○ 5 to 10 years following Parkinson's Disease diagnosis
- ○ Greater than 10 years following Parkinson's Disease diagnosis
- ○ Unsure
- ○ Prefer not to say

---

Page Break

Q13 Do you believe you have you ever experienced symptoms of any psychological conditions but not received a formal diagnosis?

*Examples may include (but are not limited to): Depression, Anxiety, Hallucinations, Delusions, Apathy, Impulsivity or compulsive behaviours, Memory problems, Dementia.*
*If completing as a carer for individual(s) with Parkinson's Disease please complete based on your knowledge of those you care for.*

- ○ Yes
- ○ No
- ○ Unsure
- ○ Prefer not to say

*Display This Question:*

*If Q13 = Yes*

*Or Q13 = Unsure*

Q14 If yes, what condition/conditions do you believe you have experienced symptoms of? Please select all that apply:

*If completing as a carer for individual(s) with Parkinson's Disease please complete based on your knowledge of those you care for.*

- ▢ Depression
- ▢ Anxiety
- ▢ Hallucinations
- ▢ Delusions
- ▢ Apathy
- ▢ Impulsivity or compulsive behaviours
- ▢ Memory problems
- ▢ Dementia
- ▢ Other, please specify: ________________________________________________
- ▢ ⊗Prefer not to say

*Display This Question:*

*If Q14 = Depression*

Q15a In a previous question you reported having experienced symptoms of depression. Approximately, when did you experience these symptoms?

*If completing as a carer for individual(s) with Parkinson's Disease please complete based on your knowledge of those you care for.*

- Before Parkinson's Disease was diagnosed
- Within the first 6 months following Parkinson's Disease diagnosis
- 6 months to 2 years following Parkinson's Disease diagnosis
- 2 to 5 years following Parkinson's Disease diagnosis
- 5 to 10 years following Parkinson's Disease diagnosis
- Greater than 10 years following Parkinson's Disease diagnosis
- Unsure
- Prefer not to say

*Display This Question:*

*If Q14 = Anxiety*

Q15b In a previous question you reported having experienced symptoms of anxiety. Approximately, when did you experience these symptoms?

*If completing as a carer for individual(s) with Parkinson's Disease please complete based on your knowledge of those you care for.*

- ○ Before Parkinson's Disease was diagnosed
- ○ Within the first 6 months following Parkinson's Disease diagnosis
- ○ 6 months to 2 years following Parkinson's Disease diagnosis
- ○ 2 to 5 years following Parkinson's Disease diagnosis
- ○ 5 to 10 years following Parkinson's Disease diagnosis
- ○ Greater than 10 years following Parkinson's Disease diagnosis
- ○ Unsure
- ○ Prefer not to say

*Display This Question:*

*If Q14 = Hallucinations*

Q15c In a previous question you reported having experienced symptoms of hallucinations. Approximately, when did you experience these symptoms?

*If completing as a carer for individual(s) with Parkinson's Disease please complete based on your knowledge of those you care for.*

- ◯ Before Parkinson's Disease was diagnosed
- ◯ Within the first 6 months following Parkinson's Disease diagnosis
- ◯ 6 months to 2 years following Parkinson's Disease diagnosis
- ◯ 2 to 5 years following Parkinson's Disease diagnosis
- ◯ 5 to 10 years following Parkinson's Disease diagnosis
- ◯ Greater than 10 years following Parkinson's Disease diagnosis
- ◯ Unsure
- ◯ Prefer not to say

*Display This Question:*

*If Q14 = Delusions*

Q15d In a previous question you reported having experienced symptoms of delusions. Approximately, when did you experience these symptoms?

*If completing as a carer for individual(s) with Parkinson's Disease please complete based on your knowledge of those you care for.*

- ○ Before Parkinson's Disease was diagnosed
- ○ Within the first 6 months following Parkinson's Disease diagnosis
- ○ 6 months to 2 years following Parkinson's Disease diagnosis
- ○ 2 to 5 years following Parkinson's Disease diagnosis
- ○ 5 to 10 years following Parkinson's Disease diagnosis
- ○ Greater than 10 years following Parkinson's Disease diagnosis
- ○ Unsure
- ○ Prefer not to say

*Display This Question:*

*If Q14 = Apathy*

Q15e In a previous question you reported having experienced symptoms of apathy. Approximately, when did you experience these symptoms?

*If completing as a carer for individual(s) with Parkinson's Disease please complete based on your knowledge of those you care for.*

- ○ Before Parkinson's Disease was diagnosed
- ○ Within the first 6 months following Parkinson's Disease diagnosis
- ○ 6 months to 2 years following Parkinson's Disease diagnosis
- ○ 2 to 5 years following Parkinson's Disease diagnosis
- ○ 5 to 10 years following Parkinson's Disease diagnosis
- ○ Greater than 10 years following Parkinson's Disease diagnosis
- ○ Unsure
- ○ Prefer not to say

*Display This Question:*

*If Q14 = Impulsivity or compulsive behaviours*

Q15f In a previous question you reported having experienced symptoms of impulsivity or compulsive behaviours. Approximately, when did you experience these symptoms?

*If completing as a carer for individual(s) with Parkinson's Disease please complete based on your knowledge of those you care for.*

◯ Before Parkinson's Disease was diagnosed

◯ Within the first 6 months following Parkinson's Disease diagnosis

◯ 6 months to 2 years following Parkinson's Disease diagnosis

◯ 2 to 5 years following Parkinson's Disease diagnosis

◯ 5 to 10 years following Parkinson's Disease diagnosis

◯ Greater than 10 years following Parkinson's Disease diagnosis

◯ Unsure

◯ Prefer not to say

*Display This Question:*

*If Q14 = Memory problems*

Q15g In a previous question you reported having experienced symptoms of memory problems. Approximately, when did you experience these symptoms?

*If completing as a carer for individual(s) with Parkinson's Disease please complete based on your knowledge of those you care for.*

- ○ Before Parkinson's Disease was diagnosed
- ○ Within the first 6 months following Parkinson's Disease diagnosis
- ○ 6 months to 2 years following Parkinson's Disease diagnosis
- ○ 2 to 5 years following Parkinson's Disease diagnosis
- ○ 5 to 10 years following Parkinson's Disease diagnosis
- ○ Greater than 10 years following Parkinson's Disease diagnosis
- ○ Unsure
- ○ Prefer not to say

*Display This Question:*

*If Q14 = Dementia*

Q15h In a previous question you reported having experienced symptoms of dementia. Approximately, when did you experience these symptoms

*If completing as a carer for individual(s) with Parkinson's Disease please complete based on your knowledge of those you care for.*

- Before Parkinson's Disease was diagnosed
- Within the first 6 months following Parkinson's Disease diagnosis
- 6 months to 2 years following Parkinson's Disease diagnosis
- 2 to 5 years following Parkinson's Disease diagnosis
- 5 to 10 years following Parkinson's Disease diagnosis
- Greater than 10 years following Parkinson's Disease diagnosis
- Unsure
- Prefer not to say

*Display This Question:*

*If Q14 = Other, please specify:*

Q15i In a previous question you reported having experienced symptoms of 'Other' psychological condition(s). Approximately, when did you experience these symptoms? If you experienced symptoms of multiple other conditions please refer to the earliest date.

*If completing as a carer for individual(s) with Parkinson's Disease please complete based on your knowledge of those you care for.*

- ◯ Before Parkinson's Disease was diagnosed
- ◯ Within the first 6 months following Parkinson's Disease diagnosis
- ◯ 6 months to 2 years following Parkinson's Disease diagnosis
- ◯ 2 to 5 years following Parkinson's Disease diagnosis
- ◯ 5 to 10 years following Parkinson's Disease diagnosis
- ◯ Greater than 10 years following Parkinson's Disease diagnosis
- ◯ Unsure
- ◯ Prefer not to say

Q16 Did you first experience physical or psychological symptoms related to Parkinson's Disease?

*If completing as a carer for individual(s) with Parkinson's Disease please complete based on your knowledge of those you care for.*

- ◯ Physical
- ◯ Psychological
- ◯ Both together
- ◯ Prefer not to say
- ◯ Unsure

**End of Block: About your Mental Health**

---

Start of Block: Impact of physical symptoms on Mental Health

Q17 Do you feel your <u>physical</u> symptoms of Parkinson's Disease impact your psychological wellbeing?

*If completing as a carer please answer based upon your perception of this for the individual(s) you care for.*

Please feel free to add additional details in the relevant box provided.

If your answer is Yes, please expand in the yes box below
If your answer is No, please outline your thoughts about the impact or how you manage to minimise the impact

○ Yes ________________________________________________

○ No ________________________________________________

○ Unsure ________________________________________________

○ Prefer not to say ________________________________________________

---

Q18 To what extent do you feel your <u>physical</u> symptoms impact on your ability to complete day to day activities?

*If completing as a carer please answer based upon your perception of this for the individual(s) you care for.*

◯ Constantly

◯ Daily

◯ Weekly

◯ Monthly

◯ Rarely

◯ Not at all

◯ Prefer not to say

---

Q19 To what extent do you feel your <u>physical</u> symptoms impact on your quality of life?

*If completing as a carer please answer based upon your perception of this for the individual(s) you care for.*

◯ Constantly

◯ Daily

◯ Weekly

◯ Monthly

◯ Rarely

◯ Not at all

◯ Prefer not to say

**End of Block: Impact of physical symptoms on Mental Health**

**Start of Block: Impact of psychological symptoms on physical wellbeing:**

Q20 Do you feel the psychological symptoms of Parkinson's Disease impact your physical functioning?

*If completing as a carer please answer based upon your perception of this for the individual(s) you care for.*

Please feel free to add additional details in the relevant box provided.

If your answer is Yes, please expand in the yes box below
If your answer is No, please outline your thoughts about the impact or how you manage to minimise the impact

○ Yes ________________________________________________

○ No ________________________________________________

○ Unsure ________________________________________________

○ Prefer not to say ________________________________________________

Q21 To what extent do you feel your <u>psychological</u> symptoms impact on your ability to complete day to day activities?

*If completing as a carer please answer based upon your perception of this for the individual(s) you care for.*

- ◯ Constantly
- ◯ Daily
- ◯ Weekly
- ◯ Monthly
- ◯ Rarely
- ◯ Not at all
- ◯ Prefer not to say

---

Q22 To what extent do you feel your <u>psychological</u> symptoms impact on your quality of life?

*If completing as a carer please answer based upon your perception of this for the individual(s) you care for.*

- ◯ Constantly
- ◯ Daily
- ◯ Weekly
- ◯ Monthly
- ◯ Rarely
- ◯ Not at all
- ◯ Prefer not to say

**End of Block: Impact of psychological symptoms on physical wellbeing:**

Start of Block: Treatments

Q23 Which treatments (if any) do you feel are beneficial for your <u>physical</u> symptoms of Parkinson's Disease? Please select all that apply.

*If completing as a carer please answer based upon your perception of this for the individual(s) you care for.*

- ☐ Medication
- ☐ Exercise
- ☐ Mindfulness
- ☐ Physiotherapy
- ☐ Occupational Therapy
- ☐ Speech and Language Therapy
- ☐ Psychology
- ☐ Psychotherapy
- ☐ Counselling
- ☐ Complementary Therapies
- ☐ ⊗Not applicable
- ☐ Other, please specify: ________________________________________________

Q24 Which treatments (if any) do you feel are beneficial for your psychological symptoms of Parkinson's Disease? Please select all that apply.

*If completing as a carer please answer based upon your perception of this for the individual(s) you care for.*

- ☐ Medication
- ☐ Exercise
- ☐ Mindfulness
- ☐ Physiotherapy
- ☐ Occupational Therapy
- ☐ Speech and Language Therapy
- ☐ Psychology
- ☐ Psychotherapy
- ☐ Counselling
- ☐ Complementary Therapies
- ☐ ⊗Not applicable
- ☐ Other, please specify: ________________________________________________

Page Break

Q25 To what extent do you feel the physical and psychological symptoms of Parkinson's Disease should be considered together by people with Parkinson's Disease and their carers?

- ○ Strongly Agree
- ○ Agree
- ○ Neither agree nor disagree
- ○ Disagree
- ○ Strongly Disagree

---

Q26 To what extent do you feel your physical and psychological symptoms of Parkinson's Disease are considered together by healthcare professionals?

*If completing as a carer please answer based upon your perception of this for the individual(s) you care for.*

- ○ Strongly Agree
- ○ Agree
- ○ Neither agree nor disagree
- ○ Disagree
- ○ Strongly Disagree

Q27 Based on your previous experience, how often have healthcare professionals discussed both the physical and psychological symptoms of Parkinson's Disease with you during the same consultation?

*If completing as a carer please answer based upon your perception of this for the individual(s) you care for.*

○ Always

○ Frequently

○ Sometimes

○ Rarely

○ Never

○ Don't Know

**End of Block: Treatments**

Start of Block: Covid-19

Q28 Do you feel Covid-19 has had an impact upon the progression of your condition?

*If completing as a carer please answer based upon your perception of this for the individual(s) you care for.*

- ○ Yes - More rapid progression
- ○ Yes - Slower progression
- ○ No
- ○ Unsure
- ○ Not applicable
- ○ Prefer not to say
- ○ Don't Know

---

Q29 Do you feel COVID-19 has impacted on the physical symptoms you experience as a result of your Parkinson's Disease?

*If completing as a carer please answer based upon your perception of this for the individual(s) you care for.*

- ○ Yes - Worsened symptoms
- ○ Yes - Improved symptoms
- ○ No
- ○ Unsure
- ○ Prefer not to say

---

Q30 Do you feel COVID-19 has impacted on the psychological symptoms you experience as a result of your Parkinson's Disease?

*If completing as a carer please answer based upon your perception of this for the individual(s) you care for.*

- ◯ Yes - Worsened symptoms
- ◯ Yes - Improved symptoms
- ◯ No
- ◯ Unsure
- ◯ Prefer not to say
- ◯ Don't Know

---

Q31 Do you feel that since legal restrictions ended that access to treatment for your physical and psychological symptoms has returned to pre-pandemic levels?

*If completing as a carer please answer based upon your perception of this for the individual(s) you care for.*

- ◯ Yes
- ◯ No
- ◯ Unsure
- ◯ Prefer not to say

*Skip To: End of Block If Q31 != No*

---

Q32 If not, what do you feel this is due to? Please select all that apply.

*If completing as a carer please answer based upon your perception of this for the individual(s) you care for.*

▢ Reduced input from therapeutic services

▢ Unable to access previous groups

▢ Natural progression of Parkinson's Disease symptoms

▢ Reduced physical activity

▢ Reduced social interaction

▢ Fear/anxiety

▢ Other, please specify: ________________________________________________

▢ ⊗Don't Know

▢ ⊗Prefer not to say

**End of Block: Covid-19**

---

Start of Block: Mattering and Anti-Mattering

Q33 Please complete the below table.

*If completing as a carer please answer based upon your feelings about your own importance.*

| | Not at all | A little | Somewhat | A lot |
|---|---|---|---|---|
| How important are you to others? | ○ | ○ | ○ | ○ |
| How much do other people pay attention to you? | ○ | ○ | ○ | ○ |
| How much would you be missed if you went away? | ○ | ○ | ○ | ○ |
| How interested are others in what you have to say? | ○ | ○ | ○ | ○ |
| How much do other people depend upon you? | ○ | ○ | ○ | ○ |

Q34 Please complete the below table.

*If completing as a carer please answer based upon your feelings about <u>your own</u> importance.*

| | Not at all | A little | Somewhat | A lot |
|---|---|---|---|---|
| How much do you feel like you don't matter? | ○ | ○ | ○ | ○ |
| How much do you feel like you will never matter to certain people? | ○ | ○ | ○ | ○ |
| How often have you been made to feel by someone that they don't care about what you think or what you have to say? | ○ | ○ | ○ | ○ |
| How often have you been treated in a way that makes you feel like you are insignificant? | ○ | ○ | ○ | ○ |
| To what extent have you been made to feel like you are invisible? | ○ | ○ | ○ | ○ |

**End of Block: Mattering and Anti-Mattering**

**Start of Block: Any further comments**

Q35 Please use the space below to add any further details you feel may be relevant regarding the potential interaction between the physical and psychological symptoms commonly experienced in Parkinson's Disease.

________________________________________________________________

________________________________________________________________

________________________________________________________________

________________________________________________________________

________________________________________________________________

**End of Block: Any further comments**

---

**End of Survey:**

We thank you for your time spent taking this survey. Your response has been recorded.

We kindly ask that you please forward the link of the survey to any other individuals you know who are living with Parkinson's Disease.
